# Predicting Future Cognitive Impairment in *De Novo* Parkinson’s Disease Using Clinical Data and Structural MRI

**DOI:** 10.1101/2021.08.13.21261662

**Authors:** Nicola Smith, Owen Williams, Lucia Ricciardi, Francesca Morgante, Thomas R Barrick, Mark Edwards, Christian Lambert

## Abstract

**BACKGROUND:** Parkinson’s disease is the second most common neurodegenerative condition and associated with increasing cognitive dysfunction as the disease progresses. However, subtle cognitive deficits can be detected at diagnosis in 42% of individuals, suggesting that damage may already be present. Our aim was to determine clinical and structural differences in those recently diagnosed with PD who later develop cognitive impairment, and whether these changes predict future cognitive decline.

**METHODS:** Clinical and imaging data was acquired from the Parkinson’s Progression Markers Initiative for 318 individuals with a diagnosis of Parkinson’s disease and baseline 3T T1-weighted MRI. The cohort was divided according to cognitive status over follow-up, with 9 individuals developing Parkinson’s disease dementia, 102 developing mild cognitive impairment and 207 remaining cognitively unaffected.

**FINDINGS:** At baseline, those who went on to develop cognitive impairment (mild cognitive impairment or dementia) were **older with more severe motor and non-motor symptoms (anosmia, rapid eye movement sleep behaviour disorder, depression)**. Grey matter loss was present in those destined for Parkinson’s disease dementia in the **precuneus, hippocampi, primary olfactory cortex, lingual gyrus, temporal cortex and cerebellum**. Those who later developed mild cognitive impairment had an attenuated but similar pattern of **grey matter loss in the temporal lobe, lingual gyrus and cerebellum**. Using support vector machines with a feature selection step, future cognitive impairment could be predicted using 11 clinical variables (AUC = 0.81), structural imaging (AUC = 0.72) or a combination of these two modalities (AUC = 0.85). These models more accurately predicted those who developed dementia (subgroup sensitivity 100%).

**INTERPRETATION:** Significant abnormalities in cortical structure is present at least three years before dementia manifests in Parkinson’s disease, with associated differences in clinical profiles. Combining this data provides a technique to accurately identify future cognitive impairment, providing a non-invasive way to stratify individuals early on.

## INTRODUCTION

Cognitive impairment is a common and disabling feature of Parkinson’s disease, particularly as the disease progresses (Compta *et al*., 2013). The prevalence of dementia in PD is 30%, with at least 75% of patients being affected after 10 years of disease (Aarsland & Kurz, 2010), causing a major impact on health and social care costs and quality of life (Soh *et al*., 2011). The spectrum of Parkinson’s disease cognitive impairment (PD-CI) can be divided in two broad categories: Parkinson’s disease with mild cognitive impairment (PD-MCI) and Parkinson’s disease with Dementia (PDD). While dementia is typically considered a feature of advanced disease (Aarsland *et al.,* 2003), evidence of PD-MCI (Litvan *et al*., 2012) can be detected in 42% of newly diagnosed PD patients (Yarnall *et al.,* 2014).

Demographic, clinical and morphological predictors of PDD have been extensively studied, mostly through cohort studies of treated Parkinson’s disease patients with long disease durations. Identified associations include older age, male gender, lower education, more severe parkinsonism, depression, Rapid eye movement sleep behaviour disorder (RBD), autonomic dysfunction, presence of MCI and visual hallucinations (Compta *et al*., 2013; Hu *et al*., 2014; Zhu *et al*., 2014). However, only a few studies prospectively assessed risk factors in newly diagnosed PD patients (Anang *et al*., 2014; Mak *et al*., 2015; Schrag *et al*., 2017). Older age appears to be the most consistent demographic risk factor in newly diagnosed Parkinson’s disease, whereas the contribution of male gender (Anang *et al*., 2014; Cholerton *et al*., 2018) and fewer years of education (Mak *et al*., 2015; Nicoletti *et al*., 2019) are less consistent. Clinical risk factors for development of MCI and dementia included presence of RBD at baseline (either assessed by polysomnography (Anang *et al*., 2014) or by RBD screening questionnaire (Schrag *et al*., 2017)), as well as autonomic dysfunction, more severe motor impairment, gait impairment and falls (Anang *et al*., 2014). Recently, studies analysing the Parkinson’s Progression Marker Initiative (PPMI) cohort have also shown an associations with low cerebrospinal fluid amyloid β42 levels, striatal region of interest dopamine transporter (DAT)-imaging uptake with the development of cognitive impairment in newly diagnosed Parkinson’s disease (Schrag *et al*., 2017; Pagano *et al*., 2018).

Widespread cortical atrophy in established PDD has been well described, particularly in medial temporal structures, posterior cingulate, precuneus, lingual gyrus and prefrontal cortex (Gee *et al*., 2017; Melzer *et al*., 2012). In contrast, the reported abnormalities in PD-MCI are variable, with a possible trend to small regions of grey matter loss in the pre-frontal and lateral-temporal cortices (Hanganu et al., 2013; Sasikumar et al., 2020; Yarnall *et al.,* 2014). More recently studies have examined the structural changes in the early phase of PD cognitive impairment (PD-CI), demonstrating significant atrophy in fronto-temporal, thalamus, accumbens and caudate compared to those without cognitive impairment (Foo *et al.,* 2017; Zhou *et al*., 2019). In a recent study using the PPMI cohort, individuals who convert to PD-MCI were found to have significantly greater right temporal pole atrophy than nonconverters at baseline (Zhou *et al*., 2019).

Notably, few neuroimaging studies have combined clinician and structural metrics to predict conversion to cognitive impairment in newly diagnosed Parkinson’s disease. Predicting cognitive impairment could potentially inform clinical discussions with patients and families regarding prognosis, could influence decisions on prescribing or selecting candidates for advanced treatments such as deep brain stimulation surgery, and help identify cohorts for clinical trials directed towards preserving cognitive function.

Our aim was to determine whether there are clinical or structural changes already present at diagnosis in individuals with Parkinson’s disease destined to develop cognitive impairment, and whether these abnormalities can be used to predict future cognitive decline. To investigate this, we used the MRI and clinical data from individuals diagnosed with Parkinson’s disease participating in the Parkinson’s Progression Markers Initiative (PPMI, Parkinson’s Progression Markers Initiative 2014).

## METHODS

### Data Acquisition and Cohort Selection

All available data were downloaded from the PPMI website (http://www.ppmi-info.org/) on the 22^nd^ November 2016. Individuals classified as “excluded”, “declined” or “pending” were removed. Sub-selecting those with a diagnosis of Parkinson’s disease, we divided the group into three categories using the most severe documented cognitive category for an individual across all time-points: Parkinson’s disease without dementia (PD-NON), PD-MCI or PDD. We removed any participant where the confidence in the cognitive category (documented by the PPMI investigators) was less than 50%. We then selected only those who had 3T T1-weighted MRI available (Tardif *et al.,* 2010). This provided a final dataset consisting of 207 PD-NON, 102 PD-MCI and 9 PDD (Fig. 1).

**Figure 1:**
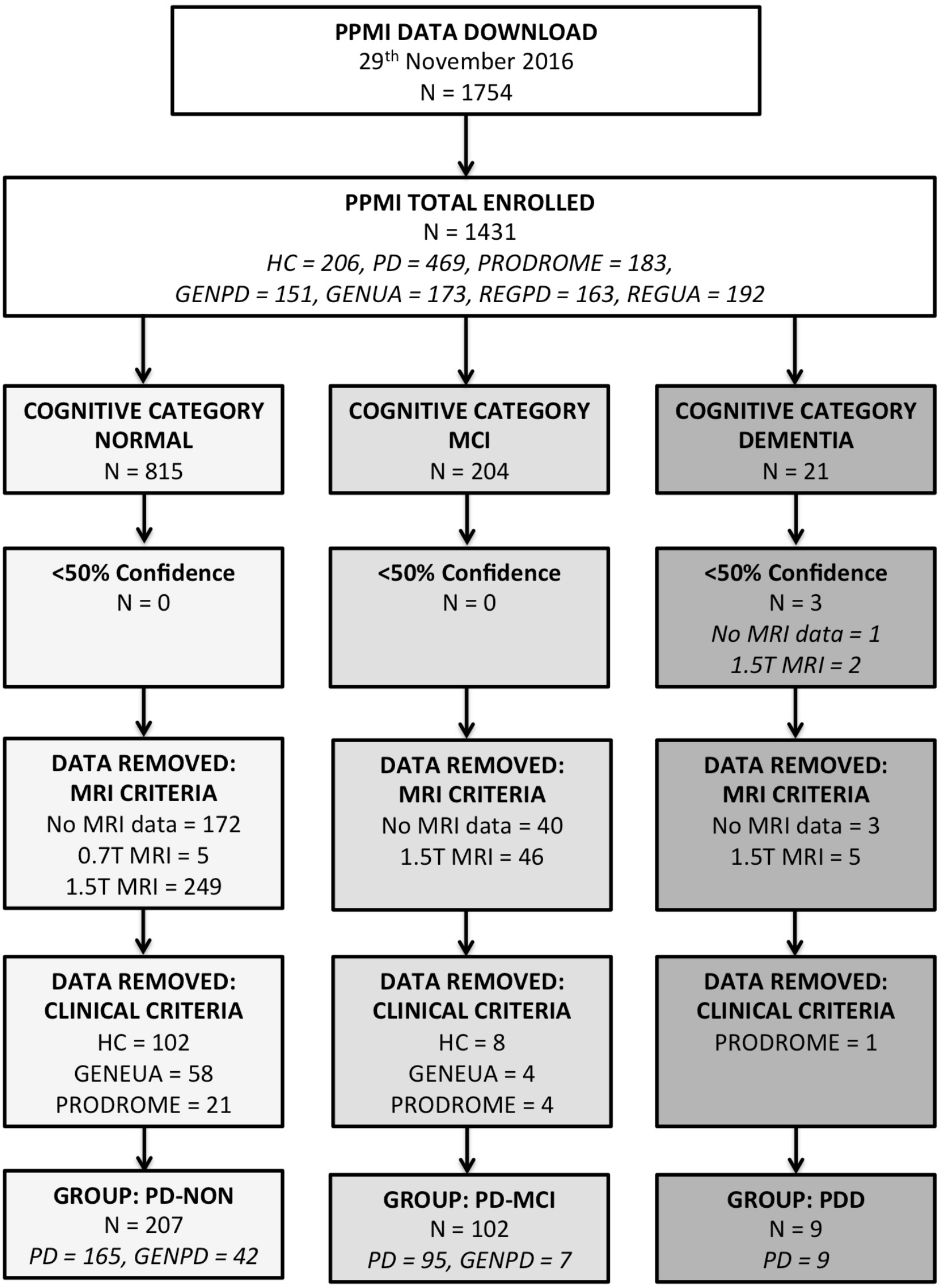
Cohort selection from the PPMI data download. Confidence level refers to PPMI documented confidence in the assigned cognitive category. *Abbreviations: HC = Healthy Control; PD = Parkinson’s disease; PRODROME = Prodromal cohort; GENPD = Genetic cohort with PD; GENUA = Genetic cohort unaffected; REGPD = Genetic Registry with PD; REGUA = Genetic registry unaffected; PD-NON = PD without cognitive impairment; PD-MCI = PD mild cognitive impairment; PDD = Parkinson’s disease dementia*.

The diagnosis of dementia or mild cognitive impairment (MCI) was defined via clinical assessment by the PPMI investigators. Full details are available on the PPMI website. The determination of dementia was based on the following criteria: (1) impaired cognitive function in more than one domain more than 1.5 standard deviations below the standardised mean, (2) decline from pre-morbid function as determined by investigator or reports from informants and (3) significant impact on daily function. The determination of MCI was based on (1) at least one cognitive domain impaired, more than one standard deviations below the standardised mean, (2) decline from pre-morbid function, and (3) lack of significant impact on daily function. Combined, the available cognitive and clinical measures provided enough evidence to satisfy the criteria for probable PDD or level 1 PD-MCI (Emre et al., 2007; Litvan *et al*., 2012). To calculate the time from diagnosis to cognitive impairment, we took the first PPMI visit where the cognitive category of interest was assigned as the time of cognitive conversion. To calculate the number of individuals who had abnormal cognitive tests at baseline, we defined abnormal as more than one standard deviation from the age-corrected norms. Single domain MCI was defined as abnormalities on two tests of one cognitive domain, and multiple-domain MCI was defined as at least one deficit measured in more than one cognitive domain.

No patient was on any dopaminergic medication at baseline. We extracted the following demographic and clinical data from the baseline PPMI visit: age, gender, years of education, date of symptom onset and date of diagnosis, Movement Disorders Society Unified Parkinson’s Disease Rating Scale (MDS-UPDRS) (Goetz *et al*., 2008) parts I-III and total (sum of parts I-III), Postural Instability and Gait Disorder (PIGD) score (Stebbins *et al*., 2013), tremor score, tremor-PIGD ratio. We also retrieved the following ratings for non-motor symptoms: REM sleep disorder questionnaire (RBDQ) (Stiasny-Kolster *et al*., 2007); Geriatric depression scale (GDS) and State-trait anxiety inventory (STAI); Epworth sleepiness scale. Autonomic function was rated by the “Scales for Outcomes in Parkinson’s disease – Autonomic” (SCOPA-AUT), and olfaction by the University of Pennsylvania Smell Identification Test (UPSIT) (Doty *et al*., 1984).

Global cognitive function was measured using the Montreal cognitive assessment (MoCA). In addition to this, the following age-norm corrected measures for four cognitive domains were also extracted: 1) Episodic memory: Hopkins verbal learning test revised (HVLT); 2) visuo-spatial function: Benton judgment of line orientation (JLO); 3) attention and executive function: Letter number sequencing (LNS) and Symbol digit modalities test (SDM). 4) Language - Semantic fluency (SF).

### Neuroimaging Pre-Processing

Using SPM12 (http://www.fil.ion.ucl.ac.uk/spm/software/spm12/), the raw DICOMS were imported and each image manually checked. The original image resolution was between 1mm and 1.1mm isotropic, so the MRI data was re-aligned to MNI orientation and re-sliced to 1mm isotropic voxel resolution in subject space (Friston *et al.,* 1995). All re-orientated T1-weighted images were segmented into grey matter (GM), white matter (WM) and cerebrospinal fluid (CSF) tissue classes (Ashburner & Friston, 2005). These segmentations were used to generate cortical thickness maps using the Voxel-Based Cortical-Thickness toolbox (Hutton *et al*., 2008) in SPM8 (this toolbox is currently incompatible with SPM12) with a sampling resolution of 0.5mm, CSF smoothness 3mm, CSF thinness 0.65 and number of dilations was set to 1.

The baseline GM, WM and CSF segmentations were then used to generate a group average template using the diffeomorphic warping “*Shoot*” algorithm in SPM12 (Ashburner & Friston, 2011). The segmentations were then warped using the resulting deformation fields, modulated with the Jacobian determinant data and smoothed with a 6mm full width half maximum (FWHM) Gaussian kernel. The CT maps were also warped using the corresponding deformation fields. The warped CT maps were then used to generate FWHM 6mm smoothed warped weighted images produced as described by Draganski et al, 2011 (Hutton *et al*., 2009; Draganski et al, 2011). Total intracranial volume (TIV) was calculated by integrating the Jacobians within the three tissue classes, with the segmentations binarised at a threshold of 0.2 (Malone *et al*., 2015).

### Statistical Analysis

All analyses were carried out using MATLAB 2016b. For each demographic and clinical variable, three group comparisons (PD-NON vs PD-MCI, PD-NON vs PDD and PD-MCI vs PDD) were assessed using the Kruskal-Wallis test, and if significant, tested further using a Mann–Whitney U test. For categorical data, the χ² test was used. Significance was set at a multiple comparison Bonferroni corrected *P* < 0.001.

For neuroimaging data, all analyses were performed using Analysis of Variance (ANOVA) in SPM12 controlling for age, handedness, total intracranial volume, gender, research centre and years of education. Peak voxel Family Wise Error (FWE) corrected *P* < 0.05 was considered significant. Significant results are displayed at both *P* < 0.001 uncorrected and FWE corrected *P* < 0.05.

### Prediction Analysis

All prediction analyses used binary SVMs in MATLAB 2016b to identify those destined for PD-CI, defined as either PD-MCI or PDD. All data classification used support vector machines and a feature selection step designed in this study to identify the most important features (detailed below and summarised in Supplementary Figure 2). Three potential feature sets were tested: Clinical data alone, MRI data alone and combined clinical and MRI data. Prediction accuracy metrics were calculated as the average over 10-fold cross validation training loops. Each of the final SVM classifiers were tested against an independent dataset that included 20% of the sample that was randomly selected from each group (40 PD-NON, 22 PD-CI consisting of 20 PD-MCI and 2 PDD). Statistical significance, using permutation testing with 1000 permutations, was set at *P* < 0.05.

The feature selection for the SVM was performed as follows (Supplementary Figure 2). Initially the complete dataset of potential features, *X*, was divided into training, *X_t_*, and independent, *X_i_*, testing datasets. A four-step process was then applied to the training data, *X_t_*, to perform feature selection and identify the best SVM classifier. At each point designated *X_t_*_’_ in steps 1-3 (Supplementary Figure 2), a random number generator was seeded using the timestamp, and the training-test splits re-allocated. The final classifier was then applied to the independent test data, *X_i_*:

1. **Feature weighting:** Using an iterative process, initially ten-fold cross validation was used and average absolute values for the feature weights (*W*) were extracted. The feature with the highest absolute weight (*W_max_*) was identified and removed from *X_t’_*. The position (i.e. relative importance) of this feature was stored in vector, *F*. This process was repeated until no potential features remained in *X_t’_*. The entire process was repeated to construct a matrix of feature order positions for different ten-fold cross validations and was used to calculate a vector containing average feature positions.
2. **Feature extraction:** Each identified feature was then sequentially added to matrix *X_t’_* according to their average position from lowest (i.e. most important) to highest (i.e. least important). At each iteration, ten-fold cross validation was used to calculate the average area under the curve of the SVM classification and this was stored in vector, *A*. The features that were selected for the prediction analysis were those that provided the maximum value in A (i.e. the best model corresponding to the maximum area under the curve).
3. **Classification of training data:** The selected features were applied in a SVM classification to the training data, *X_t’_*, in a ten-fold cross validation. Classification accuracy metrics for this final classification were calculated.
4. **Independent testing of the classifier:** The final SVM classification model was applied to the independent test data to extract independent classification accuracy metrics.

Slight adaptations to this feature identification and selection steps were required in application to the clinical data, imaging data, and combined clinical and imaging data. These are detailed below.

### Predicting Cognitive Decline: Clinical

The prediction method considered 29 clinical variables as constituents of *X* (summarised in Supplementary Material 1). To enable inclusion of the tremor/PIGD ratio, the value was converted to a standardised scale. This was achieved by normalising each of the three phenotype scores to have zero mean and standard deviation one (i.e. a z-statistic) and recombining these three rescaled scores such that values for the PIGD phenotype were between 0 and 1, indeterminate between 1 and 2 and tremor dominant between 2 and 3. Each of the 29 clinical metrics were converted to z-statistics and separated into training, *X*_t_, and test data, *X_i_*. The SVM classifier was trained on the *X_t_* as follows:

1) The SVM was trained using 10-fold cross validation, with each group containing identical proportions of PD-NON, PD-MCI and PDD. At the end of each fold, the average absolute weight vector was calculated. The feature corresponding to the maximum weight vector (i.e. contributing most to the model estimation) was removed from matrix *X_t’_*, and then the process repeated. This allowed the clinical features to be ordered according to the amount they contribute to predicting future PD-CI. To ensure a stable model, this entire process was repeated 500 times, and the order of the features set according to their average position over each permutation. 2) An SVM was trained using 10-fold cross validation, adding the reordered features one at a time and calculating the area under the receiver operator curve for each. 3) The final SVM and clinical features were selected using the one that achieved the maximum AUC, and 10-fold cross validation used to calculate the overall model accuracy metrics. 4) This final model was then applied to the independent test data, *X_i_*, and the overall accuracy metrics extracted.

### Predicting Cognitive Decline: Neuroimaging

A similar SVM technique was applied to the neuroimaging data. Initially, the group average grey matter mask was thresholded at 0.1 and used to construct a Gram matrix (*X*) from the smoothed, modulated GM voxel data over all subjects. The independent test set was removed, *X_i_*, and the SVM classifier trained on the remaining data, *X_t_*, as follows:

1) 10-fold cross validation was used. At each loop, the average weight vector was calculated. To obtain a voxel-wise representation of these weights, each feature weight was multiplied by the corresponding subjects native data vector in *X_t_*. This was repeated over all subjects, and summed to provide a final weight vector in voxel space. Voxels containing the largest absolute weight values corresponding to 0.01% of the total GM volume were then removed from *X_t_*, and a new Gram matrix calculated from the remaining data. This process was continued until the average classification error exceeded 40%. This entire process was iterated 100 times, and the order in which the voxels were removed was stored, from which an average feature map was identified consisting of 980 unique features ordered by importance. 2) An SVM was then trained using 10-fold cross validation, by including the reordered features at each loop in order of importance (high to low), and calculating the area under the receiver operator curve for the resultant SVM classification. The best (final) SVM classification was considered to include the features that achieved the maximum AUC. 3) The selected features were input to a final SVM classification and 10-fold cross validation was used to calculate the overall model accuracy metrics. 4) This final model applied to the independent test data (*X_i_*) and the independent classification accuracy metrics were extracted.

To compare the final feature map to the reported literature (PD-MCI or preclinical PD-CI (Melzer *et al*., 2012; Hanganu *et al*., 2013; Anang *et al*., 2014; Pereira *et al*., 2014; Mak et al., 2015)), we warped the group average final weight map to 1mm MNI space using FSL FNIRT. We then manually mapped the reported peak-statistical coordinates into MNI space using ITK-SNAP, creating an image. The distance between each reported statistical peak and all non-zero elements of the group average final weight map (Fig. 6) were then calculated, allowing the minimum distance to be extracted and reported.

### Predicting Cognitive Decline: Clinical and Neuroimaging Combined

The independent and training data were separated. Using the previously defined optimal features, a Gram matrix, *X_t_*, was generated for the clinical and imaging data separately, and then combined to form a matrix representing a weighted average of the two types of optimised features (i.e. clinical and imaging). To identify the optimal weighting parameters, a grid search through 100 weights between 0 and 1 was performed using the training data, *X_t’_*. At each weighting pair, the average AUC over a 10-fold cross validation was calculated (as per Step 2 above). The weighting parameters that provided the maximum AUC were selected, and then Steps (3) and (4) were then applied to determine the SVM classification accuracy metrics for *X_t_* and *X_i_*, as above.

## RESULTS

### Baseline Demographics and Clinical Profile

Out of 318 Parkinson’s disease subjects at baseline without any cognitive impairment, 102 (32.1%) developed PD-MCI and 9 (2.8%) PDD. Table 1 summarises demographical and clinical differences among the three groups at baseline visit.

**Table 1:**
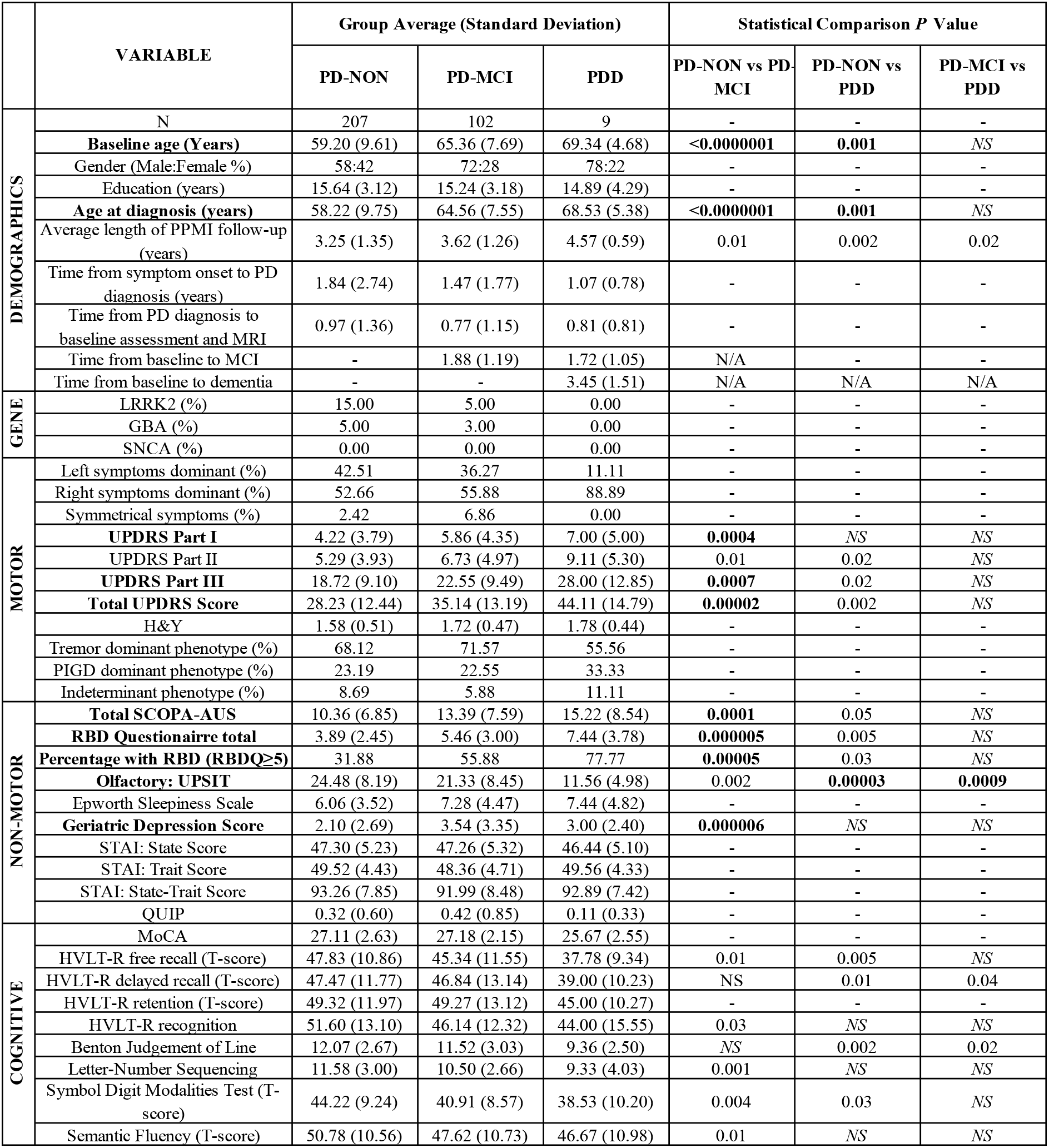
Summary of baseline demographic, clinical and psychometric parameters. Results that survive Bonferonni corrected *P* < 0.001 are highlighted in bold. NS – Not significant on Mann-Whitney U test (*P* < 0.05). Dash indicates not significant on Krushkal-Wallis test (or chi squared test for categorical descriptors).

Years of education and gender distribution were comparable among the three groups. At diagnosis, PD-MCI and PDD were significantly older compared to PD-NON. There were no significant group differences between time of symptom onset to PD diagnosis. The PDD group did have a longer amount of follow-up in the PPMI study (average 4.57 years). Nevertheless, there were no significant differences in the time to MCI conversion between PDD and PD-MCI groups; moreover, the time to PDD conversion (3.45y) was less than the average amount of follow-up in the PD-MCI group (3.62y). This suggests that the disease progression in the PDD group is more aggressive, rather than a consequence of longer follow-up in the PPMI, and is reflected by the five-year progression of their clinical variables (Fig. 2).

**Figure 2:**
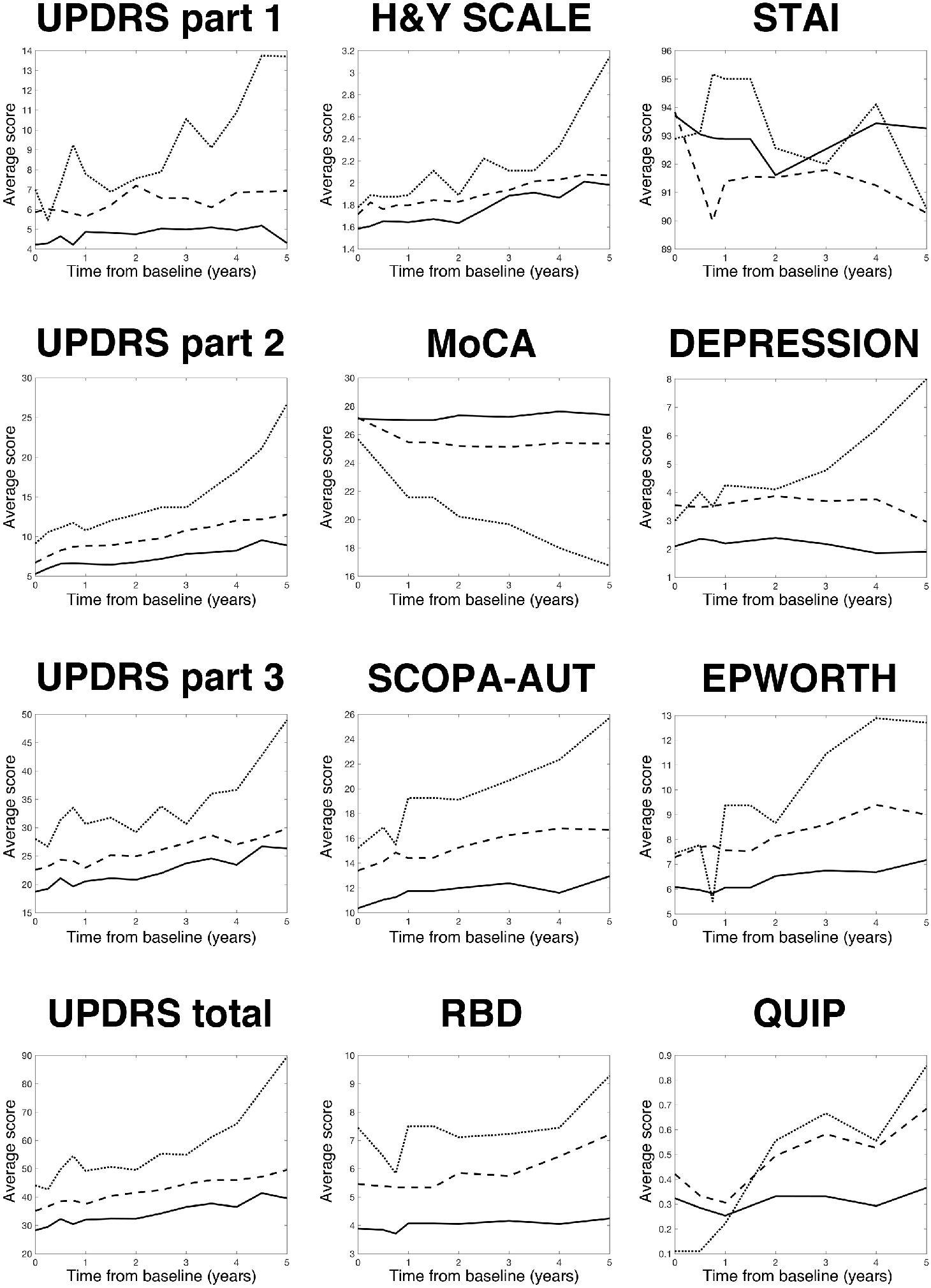
Longitudinal progression in motor and non-motor test scores over five years of follow-up. Note the average time to MCI and PDD was 1.8y and 3.45y respectively. Solid line = PD-NON, dash line = PD-MCI, dotted line = PDD. *Abbreviations: UPDRS – Unified Parkinsons Disease Rating Scale (MDS version); H&Y = Hoehn and Yahr; MoCA – Montreal Cognitive Assessment; SCOPA-AUT-Scales for Outcomes in Parkinson’s disease – Autonomic; RBD – Rapid eye movement behavioural disorder questionnaire; STAI - State- Trait Anxiety Inventory; Depression - Geriatric Depression Scale; Epworth – Epworth Sleepiness Scale; QUIP - Questionnaire for Impulsive-Compulsive Disorder in Parkinson’s Disease*.

Groups destined for cognitive impairment (either PD-MCI or PDD) had worse motor symptoms as per MDS-UPDRS III, and more severe non-motor features in the form of hyposmia, RBDQ score, depression and autonomic symptoms compared to the PD-NON group. Between the groups who later develop cognitive impairment, those destined for PDD had significantly worse hyposmia compared to PD-MCI.

On average, we found at least one abnormal cognitive test in 18.10% of all PD patients at baseline (Table 2). Patients who developed PD-MCI and PDD at follow-up had more abnormal scores that crossed different cognitive domains at baseline compared to PD-NON (*P* < 0.001, Supplementary Table 1). However there were no differences in any of the individual cognitive scores that survived Bonferroni correction (Table 1).

**Table 2:** **Full cognitive profile data at baseline** showing the percentage of individuals with abnormal scores.

|  | Percentage of cohort with abnormal*<br>baseline scores (%) |  |  |
| --- | --- | --- | --- |
| Final cognitive diagnosis | PD-NON | PD-MCI | PDD |
| MoCA | 20.3 | 18.6 | 30.0 |
| HVLT-R free recall | 21.7 | 29.4 | 66.7 |
| HVLT-R delayed recall | 20.8 | 27.5 | 55.6 |
| HVLT-R retention | 16.4 | 18.6 | 30.0 |
| HVLT-R recognition<br>discrimination | 16.4 | 34.3 | 30.0 |
| Benton Judgement of Line<br>Orientation Total | 7.72 | 15.69 | 44.4 |
| Letter-Number Sequencing | 4.3 | 7.8 | 30.0 |
| Symbol Digit Modalities Test | 22.7 | 23.5 | 55.6 |
| Semantic Fluency | 9.1 | 18.6 | 11.1 |

### Baseline Voxel-Based Morphometry

Widespread regions of reduced GM density were found in those destined for cognitive impairment that appears to form a progressive spectrum of atrophy between PD-NON, PD-MCI and PDD (Fig. 3). Comparing PD-NON and PD-MCI, the latter group was associated with reduced GM in the cerebellar association zone (Crus I/II), the left angular gyrus, left ventral pre-motor cortex and lingual gyrus. Progressing from PD-MCI to PDD, revealed additional areas of reduced GM bilaterally in the medial precuneus, posterior cingulate, hippocampi, primary olfactory cortex, and cerebellum (lobules VI, VIII). Comparing PD-NON to PDD demonstrates the full extent of the atrophy in PDD. Whilst additional areas are seen between PD-NON versus PDD that do not appear in the PD-MCI analyses, these may represent regions of evolving atrophy in PD-MCI that fall midway between the PD-NON and PDD spectrum, but are not sufficiently different to achieve significance in either comparison.

**Figure 3:**
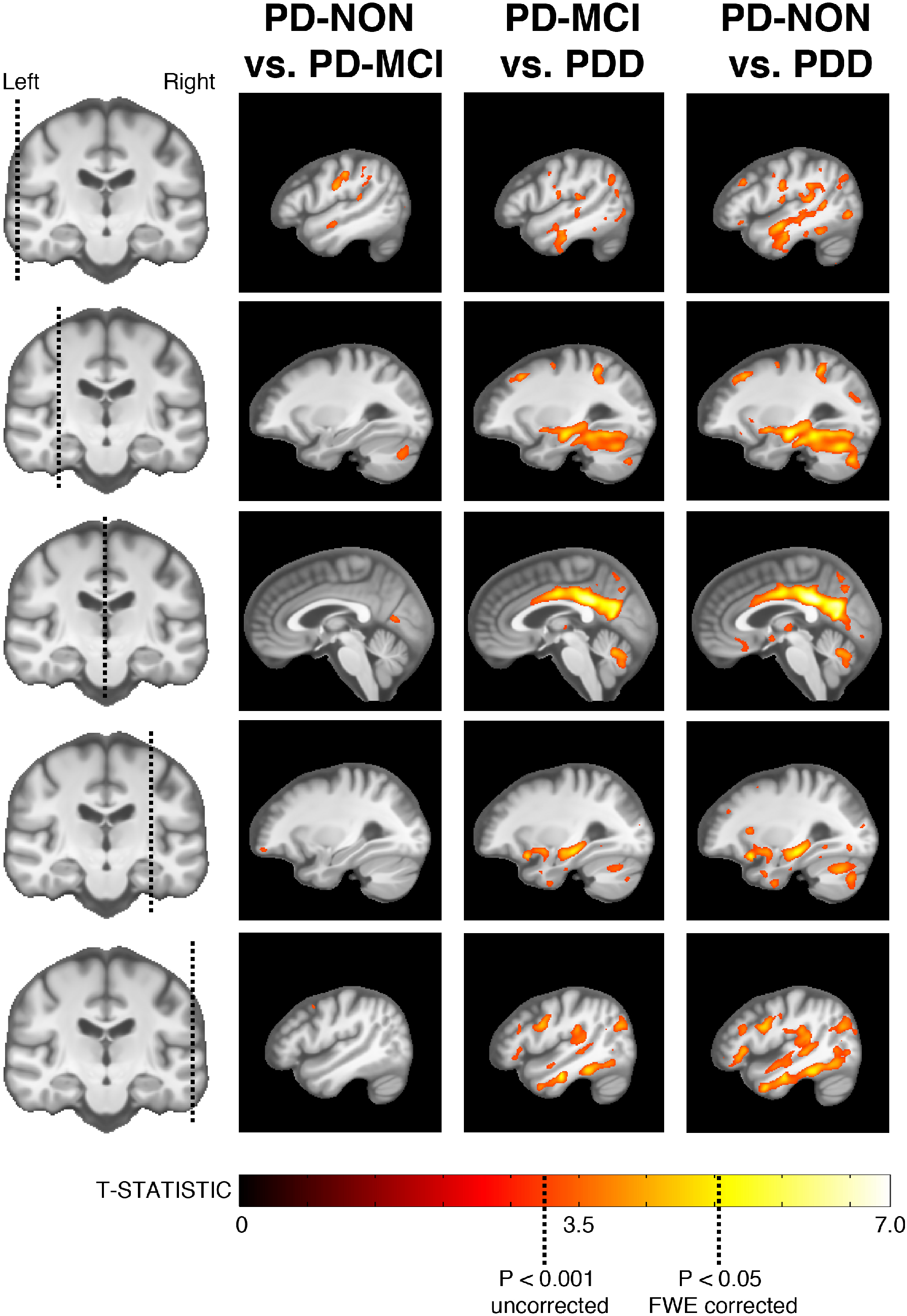
Grey matter voxel-based morphometry. Significant regions of reduced grey matter density in PD subjects with preclinical cognitive impairment (MCI or dementia).

No regions of reduced WM were observed in PD-MCI compared to PD-NON. Reduced WM in PDD was observed compared to PD-MCI in the fronto-parietal WM bilaterally, the isthmus of the corpus callosum and the anterior temporal WM (Fig. 4).

**Figure 4:**
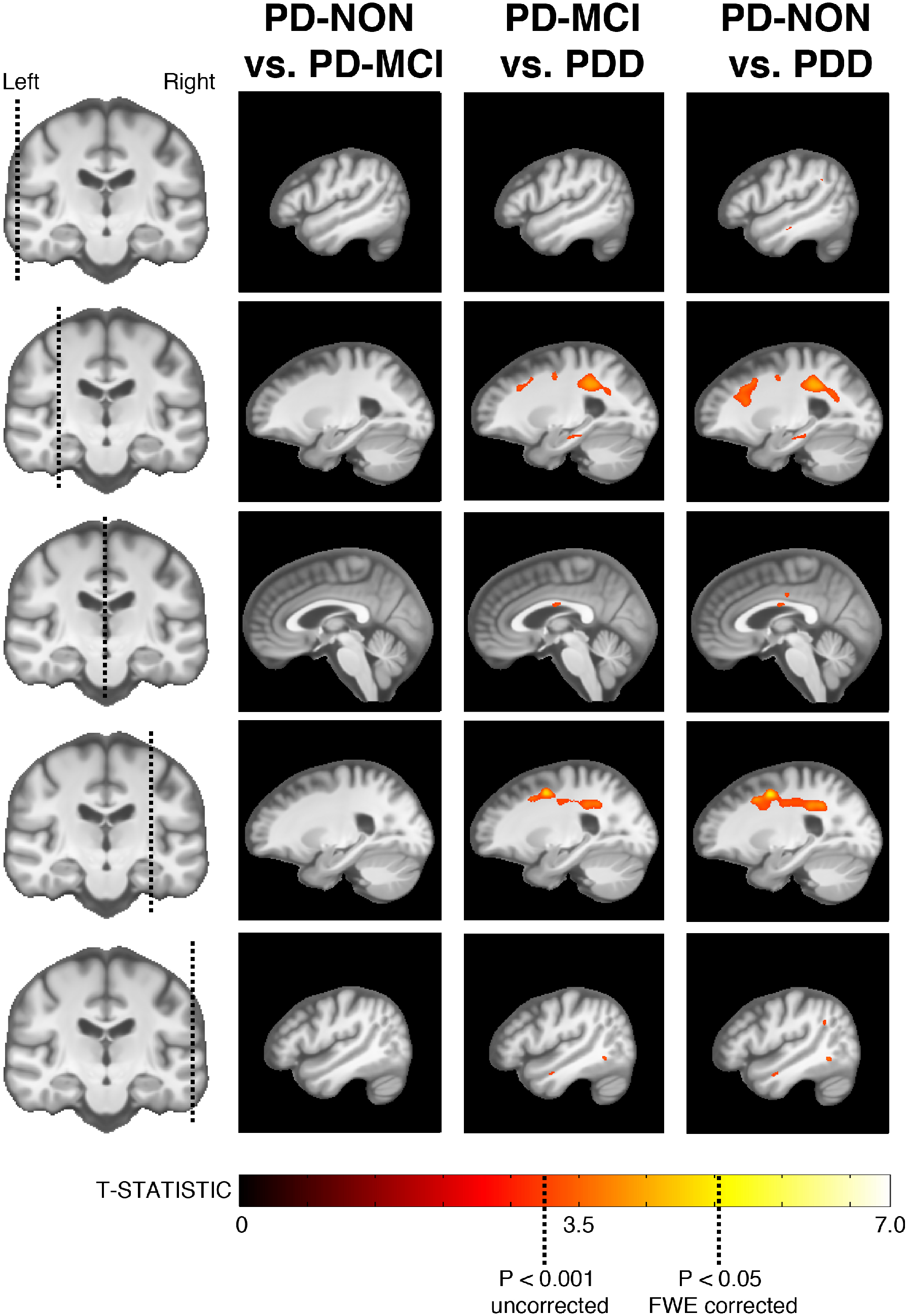
White matter voxel-based morphometry. Significant regions of reduced white matter density in PD subjects with preclinical cognitive impairment (MCI or dementia).

Voxel-based cortical thickness was also evaluated between the three groups at the time of PD diagnosis. No regions survived the FWE corrected *P* < 0.05. Given previous reports of differences (Mak *et al*., 2015), we inspected *P* < 0.001 uncorrected. This revealed several regions of thinning that were similar to the VBM analysis (Supplementary Figure 1).

### Predictors of Cognitive Decline: Clinical Features

The optimal SVM model consisted of 11 clinical variables (Fig. 5a and 5b) which were: Age at Parkinson’s disease diagnosis, HVLT-R recognition discrimination index, gender, letter number sequencing, SCOPA-AUT, RBDQ total, GDS, HVLT-R total recall, MoCA total, Epworth sleepiness scale, and symbol digit modalities.

**Figure 5:**
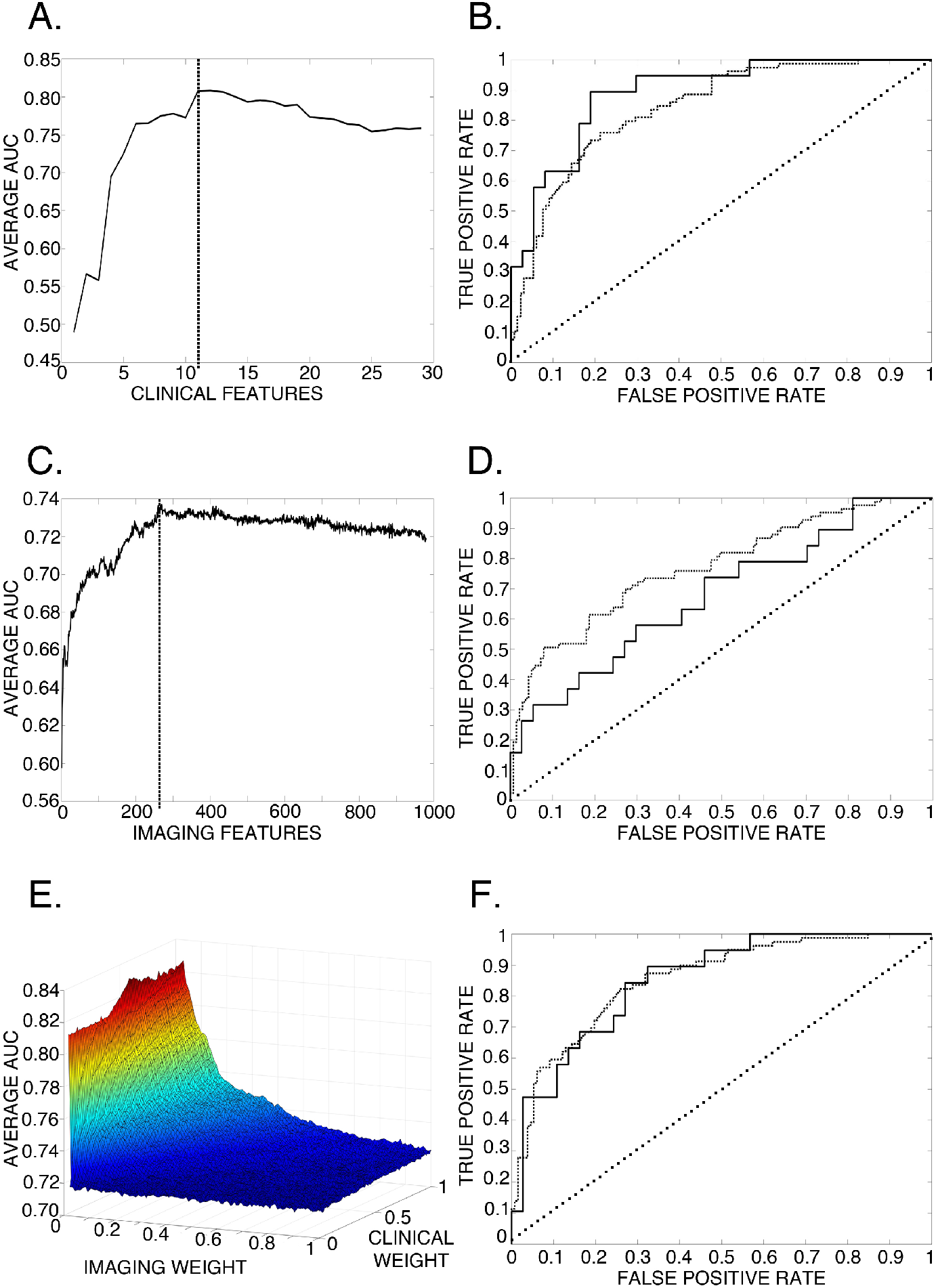
SVM results. A - Clinical optimisation, with 11 features providing the maximum average AUC. B - Receiver Operator Curve (ROC) for clinical predictions. The dashed line indicates the ROC for the model (from training data) while the solid line indicates the ROC for the independent test data. C - Imaging optimisation, with 269 features providing the maximum average AUC (see Figure 6 for the feature map). D - Receiver Operator Curve for imaging predictions. The dashed line indicates the ROC for the model (from training data) while the solid line indicates the ROC for the independent test data. E - Combined clinical-imaging grid search to define weighting parameters to produce weighted-average kernel for the analysis of clinical and neuroimaging data. F - Receiver Operator Curve for the combination of clinical and neuroimaging data. The dashed line indicates the ROC for the model (from training data) while the solid line indicates the ROC for the independent test data.

The average classification metrics (Supplementary Table 2) were: balanced prediction accuracy 73%, AUC 0.81, sensitivity 62%, specificity 85%, positive predictive value 77% and negative predictive value 76%. Applying the final model to independent test data resulted in a balanced accuracy of 76%, AUC 0.89, sensitivity 63%, specificity 88%, positive predictive value 75% and negative predictive value 82% (permutation testing *P <* 0.001). These results imply that our clinical model can significantly predict future cognitive impairment, and that this generalises well to previously unseen data.

Despite the small numbers of PDD, all training loops were balanced allowing prediction accuracies for the three cognitive sub-groups to be extracted. Over the entire cross validation, those with positive predictions were more likely to ultimately develop PDD (sensitivity 100%), compared to PD-MCI (sensitivity 53%).

### Predictors of Cognitive Decline: Neuroimaging Features

The optimal MRI SVM model consisted of 269 features (Fig. 5c and 5d), corresponding to 27% of all available grey matter. The distribution of these features overlap with VBM findings, but included more of the cerebellum and dorso-lateral temporal lobe, and additional regions such as the bilateral striatum, orbitofrontal cortex and entorhinal cortex (Fig. 6). This map corresponds to the previously reported abnormalities in PD-MCI or preclinical PD-CI, with 31% of published peak voxel coordinates completely overlapping and the remainder within 4.72mm (+/-3.26mm).

**Figure 6:**
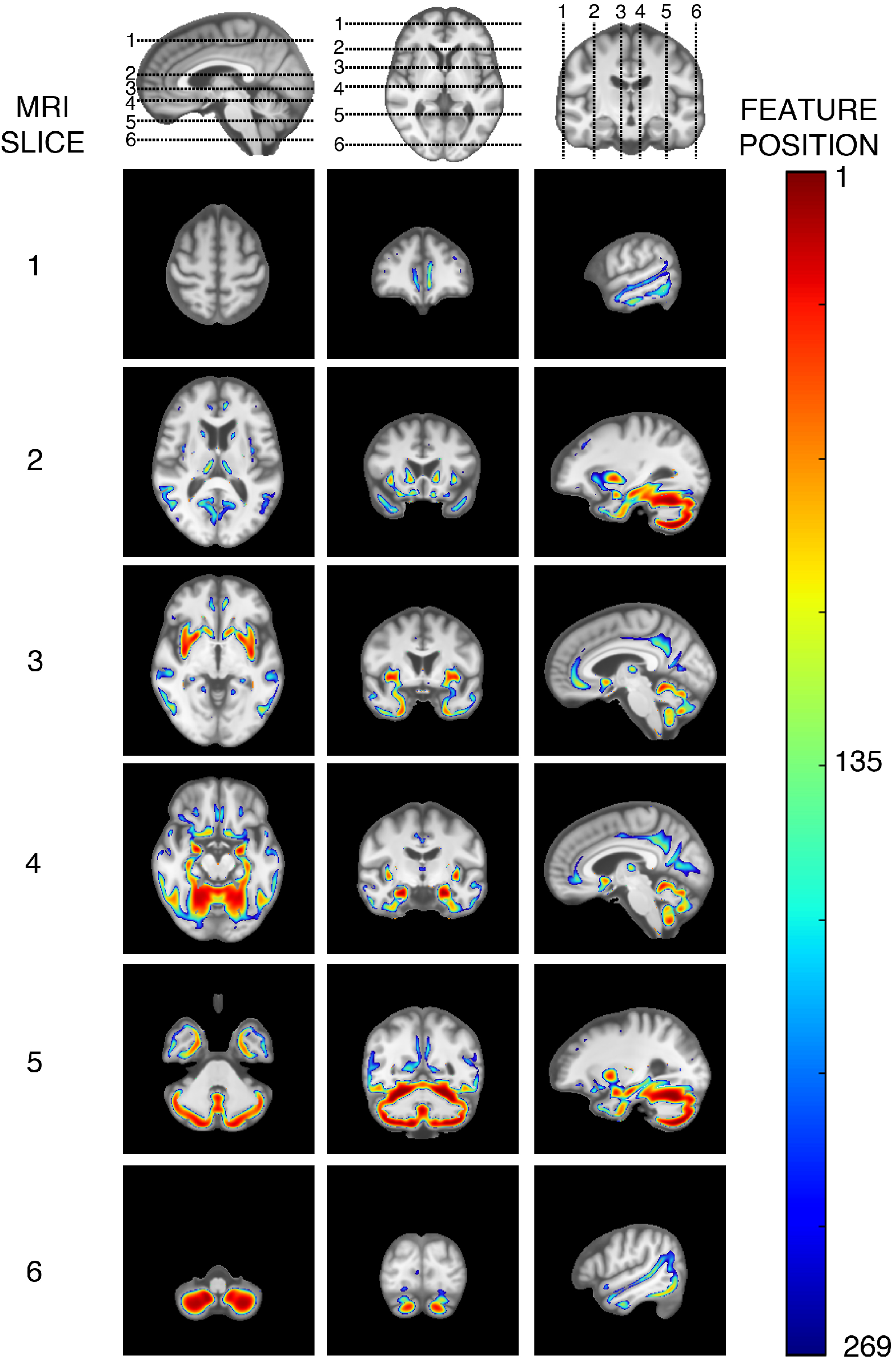
Spatial distribution of the final 269 imaging features contributing to the optimal SVM predictions. The ordering of the features has been preserved, corresponding to the largest (1) to smallest (269) SVM weights. Whilst these correspond with the voxel-based morphometry results, there is greater representation of the cerebellum and dorso-lateral temporal lobe, and the addition of regions such as the bilateral striatum, orbitofrontal cortex and entorhinal cortex.

The average classification metrics (Supplementary Table 2) were: balanced prediction accuracy 69%, AUC 0.72, sensitivity 46%, specificity 93%, positive predictive value 82% and negative predictive value 71%. Applying the final model to independent test data resulted in a balanced accuracy of 61%, AUC 0.66, sensitivity 37%, specificity 86%, positive predictive value 58% and negative predictive value 71% (permutation testing *P* < 0.01). These results imply that the imaging model can also significantly predict future cognitive impairment, though not as accurately as clinical data alone. Again, the PDD subgroup was more accurately classified compared to PD-MCI (balanced accuracy 91% vs. balanced accuracy 65%). These results highlight that whilst cognitive impairment in PD is a heterogeneous entity, there is a distinct subgroup that will progress to dementia early on, with no differences in disease duration, average time to MCI or baseline cognitive profile, but with profound abnormalities in brain structure during the pre-symptomatic phase.

### Predicting Cognitive Decline: Combined Clinical-Neuroimaging Features

Finally, we combined the clinical and imaging parameters. Using a grid search, the optimal model had a clinical weighting of 0.53 and imaging weighting of 0.01 (Fig. 5e and 5f). The average classification metrics (Supplementary Table 2) using the combined clinical imaging data were: balanced prediction accuracy 74%, AUC 0.85, sensitivity 62%, specificity 87%, positive predictive value 75% and negative predictive value 76% (permutation testing *P <* 0.001). Applying the final model to the independent test data resulted in a balanced accuracy of 78%, AUC 0.86, sensitivity 63%, specificity 92%, positive predictive value 80% and negative predictive value 82%. These results show that the combined model is the most accurate, and generalises to unseen data. Again, positive predictions were more likely in PDD (sensitivity 100%), compared to PD-MCI (sensitivity 53%). Combination of clinical and neuroimaging features in the model provides positive prediction probabilities for developing PDD, PD-MCI and PD-NON of 0.61, 0.32 and 0.08 respectively. Whilst the clinical data is the major contributor to this (Fig. 5e) the additional MRI data improves the specificity by better discriminating the PD-NON cohort.

## DISCUSSION

This work demonstrates that extensive grey and white matter loss is present at least three years before cognitive impairment clinically manifests in Parkinson’s disease. Moreover, the structural abnormalities in newly diagnosed Parkinson’s disease who later develop PD-MCI overlap with those encountered in the group developing PDD suggesting an intermediate stage. Those who develop PD-MCI or PDD are significantly older with more severe motor disability, hyposmia, depression and higher frequency of RBD symptoms at baseline. Those destined for early PDD compared to PD-MCI have more severe hyposmia. Finally, we show that future cognitive impairment in Parkinson’s disease can be predicted using clinical variables (AUC = 0.81), structural imaging (AUC = 0.72) or a combination of these two modalities (AUC = 0.85), and that these models accurately predict the emergence of early PDD (sensitivity 100%). These results suggest there is a distinct subgroup that experience earlier cognitive decline and are associated with a characteristic pattern of brain atrophy.

Parkinson’s disease is diagnosed once the motor symptoms become clinically detectable. This transition occurs once 60-70% of the dopaminergic neurons in the substantia nigra have been lost, and is the hallmark of stage III disease (Braak *et al*., 2003). In our study, there was no difference between motor symptom onset to diagnosis and baseline assessment in the three groups, suggesting that they passed this threshold at a similar time. Following a diagnosis of Parkinson’s disease, there are three main pathological entities that can interact or occur in isolation which may contribute to the development of dementia: Lewy-body disease, Alzheimer’s pathology and cerebrovascular damage (Hely *et al*., 2008; Melzer et al., 2012; Shimada *et al*., 2013; Gomperts *et* al., 2016; Irwin *et al*., 2017). Here, we have shown that a spectrum of atrophy is present at diagnosis and can predict future cognitive decline, particularly in the cerebellum, temporo-parietal cortex and primary olfactory structures. The grey matter loss detected in the group destined for PDD corresponded well with regions of atrophy in established PDD (Melzer *et al*., 2012) but with much less involvement of the frontal regions and the striatum. Additionally, regions of reduced grey matter in the cerebellum, precuneus and posterior cingulate overlapped with abnormalities found in patients with clinical dementia with Lewy bodies or PDD who also have evidence of Aβ Alzheimer’s pathology on PET amyloid imaging (Shimada *et al*., 2013). It may be that this particular sub-group, who rapidly decline cognitively and are known to progress more rapidly to death (Hely *et al*., 2008), contain a significant proportion of individuals with co-existing Alzheimer’s pathology, a hypothesis which is consistent with the histological data (Irwin *et al*., 2017).

The PD-MCI group displayed an attenuated but similar pattern of grey matter loss in the temporal lobe, lingual gyrus and cerebellum. This may represent an intermediate form between PD-NON and PDD. The SVM feature map represents regions where damage can jointly contribute to the emergence of PD-CI irrespective of the cause, and involved areas known to be affected by dementia with Lewy bodies, Alzheimer’s disease and PDD (Colloby *et al*., 2014) (Fig. 6). This may account for variation seen in previous studies where the precise results will depend upon the mixture of pathologies sampled by that particular cohort.

Recently, Schrag and colleagues used the PPMI dataset to develop a model to predict future cognitive impairment in recently diagnosed *de-novo* Parkinson’s disease (Schrag *et al*., 2017) followed up for 2 years. They found the five most strongly associated variables with cognitive impairment were age, UPSIT score, RBDQ score, CSF Aβ42, and mean caudate DAT uptake. Combining these with logistic regression gave an AUC of 0.80 for predicting future cognitive impairment. The PPMI cohort analysed in our study have a longer follow-up (up to 5 years), which allowed us to include a higher proportion of patients with cognitive impairment. We confirmed that baseline age, UPSIT score, GDS and RBDQ scores are significantly different in those destined for cognitive decline, but also show differences between SCOPA-AUT in agreement with previous work (Anang *et al*., 2014). Additionally, we have achieved a model that can more accurately predict future cognitive decline (AUC 0.85) and correctly classifies 100% destined for PDD. Whilst the differences in prediction accuracies may partly be due to greater numbers of PD-MCI or PDD in this work, the major advantage of our approach is it only requires non-invasive data. Despite the MRI component only making a minor contribution to the final model in this work, it did make a significant difference. Additionally, this work uses an extremely simple imaging modality and methodological approach, and it is likely that multimodal, quantitative MRI measures may further improve these by better detecting microstructural brain abnormalities at an individual subject level (Lambert *et al*., 2013). However, future work is required to acquire these high quality quantitative datasets for this patient cohort.

The small group size of the Parkinson’s disease dementia cohort is a limitation to further sub-group analysis in this work. Additionally, the cognitive profiles that could only provide level one evidence of PD-MCI or PDD, which would contribute to the clinical heterogeneity and may well account for the lower specificity in predicting PD-MCI. When this analysis was undertaken, the genetic cohort arm of the PPMI was only 54% complete, and none had been followed up for more than three years, which will account for the low proportions of SNCA or GBA mutations (Schapira 2015) associated with cognitive impairment in this work. The MRI data collected via the PPMI combines a variety of imaging resolutions, scanner manufacturers and MRI field strengths, hence we chose to analyse a more homogenous 3T cohort to avoid a major source of non-experimental error (Tardif *et al.,* 2010). Finally, this work used a relatively simple classifier and possibly a combination of quantitative multimodal imaging, detailed clinical phenotyping and more advanced classification methods may improve classification parameters further.

## CONCLUSION

In summary, this work demonstrates significant abnormalities in cortical structure at least three years before dementia clinically manifests in Parkinson’s disease and demonstrates that future cognitive impairment in Parkinson’s disease can be accurately predicted using non-invasive baseline data. This provides a quick and simple technique to stratify individuals with PD early in the disease course.

## Data Availability

Data from the Progression in Parkinson's Markers Initiative was used for this work

## Acknowledgements

Data used in the preparation of this article were obtained from the Parkinson’s Progression Markers Initiative (PPMI) database (www.ppmi-info.org/data). For up-to-date information on the study, visit www.ppmi-info.org. PPMI – a public-private partnership – is funded by the Michael J. Fox Foundation for Parkinson’s Research and funding partners, including AbbVie, Avid, Biogen, Bristol-Myers Squibb, Covance, GE Healthcare, Genentech, GlaxoSmithKline, Lilly, Lundbeck, Merck, Meso Scale Discovery, Pfizer, Piramal, Roche, Servier, Teva, and UCB.

## SUPPLEMENTARY MATERIAL

**Supplementary Figure 1:**
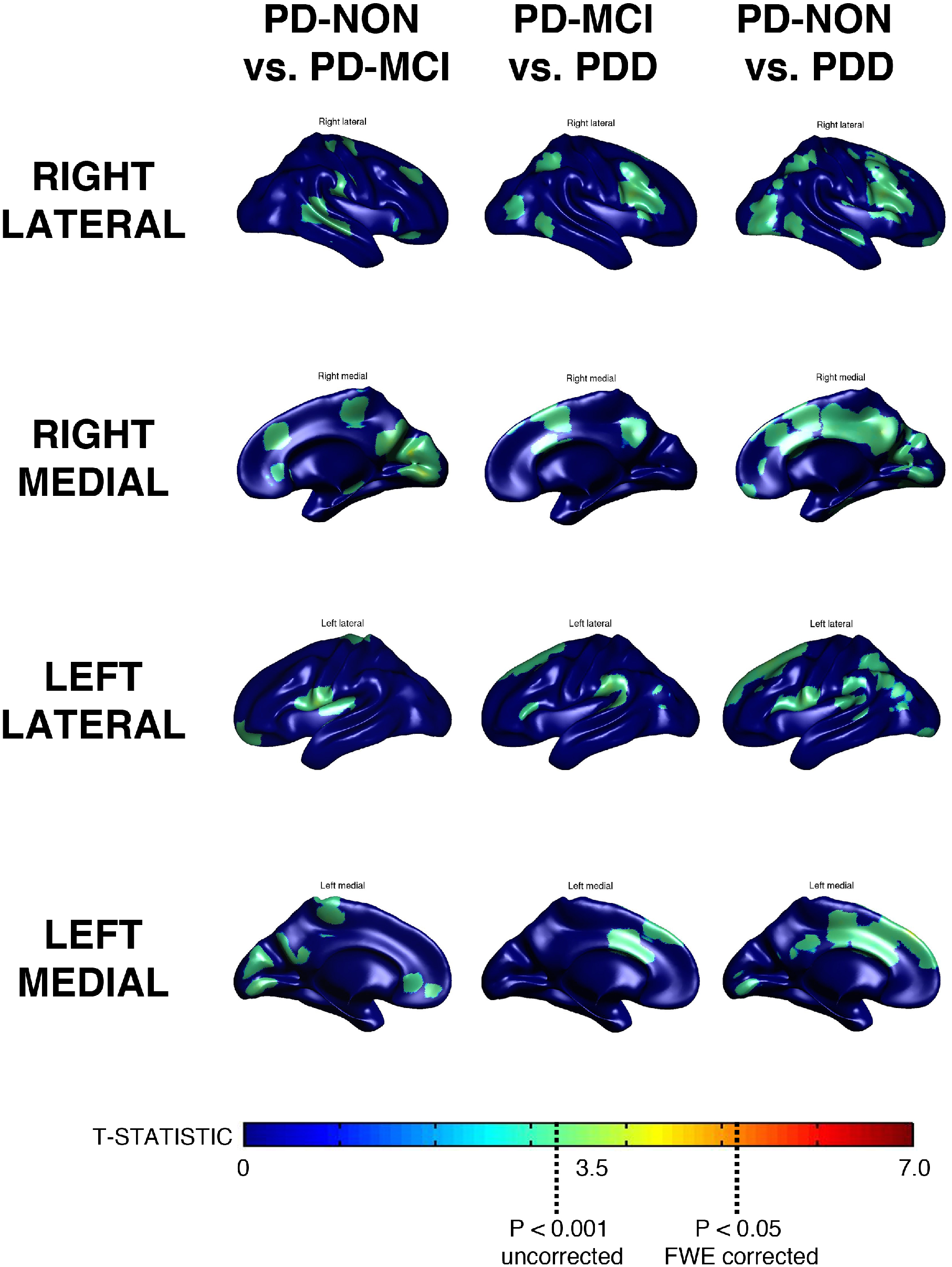
Cortical thickness. Regions of reduced cortical thickness (P<0.001) in PD subjects with preclinical cognitive impairment (MCI or dementia).

**Supplementary Figure 2:**
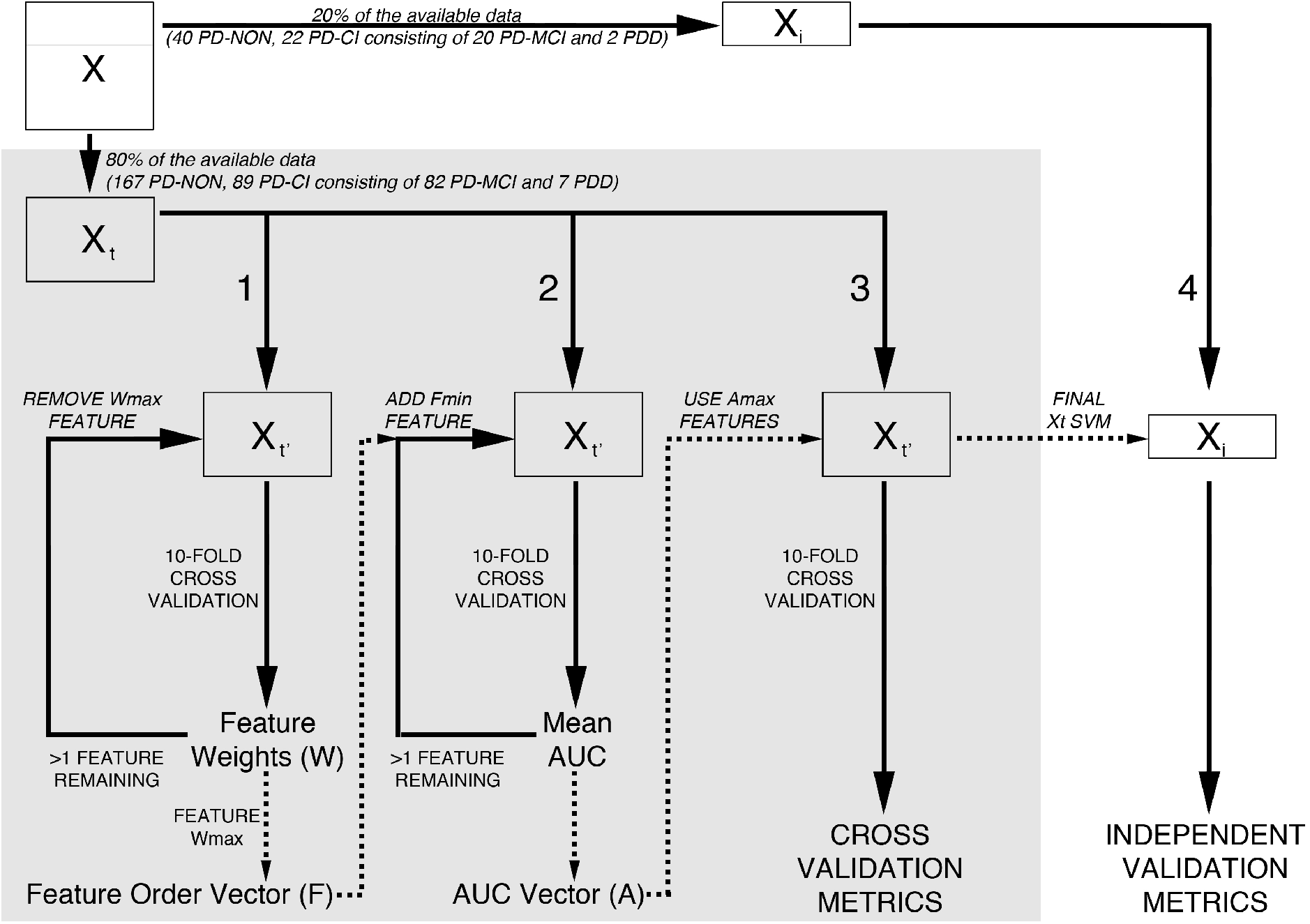
Summary of Support Vector Machine training and feature selection pipeline. The full dataset, *X*, is divided into training, *X_t_*, and independent, *X_i_*, test datasets. At each point *X_t_*_’_ in steps 1-3, a random number generator is seeded using the timestamp, and the training-test splits re-allocated. The following is performed on the training data, *X_t_*: 1) Using an iterative process, ten-fold cross validation is used to obtain average absolute values for the feature weights, *W*. The feature with the highest weight, *W_max_*, is identified and removed from *X_t_* and the position (i.e. the relative importance) of this feature is stored. This process is repeated until no features remain. Step 1) is then repeated to construct a matrix of feature positions from which the average feature location vector, *F*, is calculated. 2) The identified features are then sequentially added to create matrix *X_t’_* according to their average position from lowest (i.e. most important) to highest (i.e. least important). At each iteration, ten-fold cross validation is used to calculate the average area under the curve which is stored in vector *A*. 3) The features that correspond to the maximum value in *A* (i.e. the best model) are then applied in a ten-fold cross validation of the test dataset, *X_t_*, to calculate the overall model accuracy metrics for the training data. 4) The same features are applied to the independent test data, *X_i_*, and the accuracy metrics extracted. Note at each point, *X_t_*_’_, in steps 1-3, a random number is used to assign subjects within each 10-fold cross validation.

**Supplementary Table 1:** Percentage of individuals at baseline with abnormal test scores affecting single or multiple domains, using the definitions provided by the MDS-Task Force on PD-MCI.

|  |  | BASELINE COGNITIVE CLASSIFICATION<br>(Percentage of Cohort) |  |  |
| --- | --- | --- | --- | --- |
|  |  | NORMAL | SINGLE DOMAIN<br>DEFICIT | MULTIPLE<br>DOMAIN DEFICIT |
| COHORT | PD-NON | 59.91 | 14.00 | 26.09 |
|  | PD-MCI | 39.22 | 16.67 | 44.11 |
|  | PDD | 11.11 | 22.22 | 66.67 |

**Supplementary Table 2:** SVM accuracy metrics, top is the average over 10-fold cross-validation, bottom is the independent training set. Standard deviations provided in brackets. *Significantly higher P<0.05 compared to imaging alone. **Significantly higher compared to clinical alone.

| **10-FOLD CROSS VALIDATION AVERAGE** |  |  |  |
| --- | --- | --- | --- |
|  | **CLINICAL** | **IMAGING** | **JOINT** |
| **AUC** | 0.81 (0.11) | 0.72 (0.13) | 0.85 (0.09)* |
| **Balanced Accuracy (%)** | 73 (0.09)* | 69 (0.08) | 74 (0.07)* |
| **Sensitivity (%)** | 62 (13)* | 46 (13) | 62 (12)* |
| **Specificity (%)** | 85 (12) | 93 (6)** | 87 (6) |
| **PPV (%)** | 77 (16) | 82 (14) | 75 (9) |
| **NPV (%)** | 76 (7) | 71 (6) | 76 (6) |
| **INDEPENDENT TEST DATA** |  |  |  |
|  | **CLINICAL** | **IMAGING** | **JOINT** |
| **AUC** | 0.89 | 0.66 | 0.86 |
| **Balanced Accuracy (%)** | 76 | 61 | 78 |
| **Sensitivity (%)** | 63 | 37 | 63 |
| **Specificity (%)** | 88 | 86 | 92 |
| **PPV (%)** | 75 | 58 | 80 |
| **NPV (%)** | 82 | 71 | 82 |

**Supplementary Material 1:** Baseline clinical variables used for building the SVM model

## SUPPLEMENTARY MATERIAL 1

Baseline clinical variables used for building the SVM model

1. Baseline age
2. Gender
3. Years of education
4. UPDRS part 1 score
5. UPDRS part 2 score
6. UPDRS part 3 score
7. UPDRS total score
8. Hoehn and Yahr scale
9. Tremor score
10. PIGD score
11. TREM/PIGD ratio (normalised to a continuous scale as described in the methods)
12. SCOPA-AUT score
13. RBD questionnaire score
14. UPSIT score
15. Epworth score
16. Geriatric depression scale
17. State-trait anxiety inventory - State component
18. State-trait anxiety inventory - Trait component
19. State-trait anxiety inventory - Total score
20. QUIP total score
21. MoCA total score
22. Hopkins verbal learning test revised - Total recall (T-score)
23. Hopkins verbal learning test revised - Delayed recall (T-score)
24. Hopkins verbal learning test revised - Retention (T-score)
25. Hopkins verbal learning test revised - Recognition discrimination index (T-score)
26. Benton judgment of line orientation (age and education scaled score)
27. Letter number sequence (scaled score)
28. Symbol digit modalities test (T-score)
29. Semantic fluency (scaled score)

